# Digital reconstruction of infraslow activity in human intracranial ictal recordings using a deconvolution-based inverse filter

**DOI:** 10.1101/2022.06.20.22276557

**Authors:** Somin Lee, Julia Henry, Andrew K. Tryba, Yasar Esengul, Peter Warnke, Shasha Wu, Wim van Drongelen

## Abstract

Infraslow activity (ISA) is a biomarker that has recently become of interest in the characterization of seizure recordings. Recent data from a small number of studies have suggested that the epileptogenic zone may be identified by the presence of ISA. Investigation of low frequency activity in clinical seizure recordings, however, has been hampered by technical limitations. EEG systems necessarily include a high-pass filter early in the measurement chain to remove large artifactual drifts that can saturate recording elements such as the amplifier. This filter unfortunately attenuates legitimately seizure-related low frequencies, making ISA difficult to study in clinical EEG recordings. In this study, we present a deconvolution-based digital inverse filter that allows recovery of attenuated low frequency activity in intracranial recordings of temporal lobe epilepsy patients. First, we show that the unit impulse response (UIR) of an EEG system can be characterized by differentiation of the system’s step response. As proof of method, we present several examples that show that the low frequency component of a high-pass filtered signal can be restored by deconvolution with the UIR. We then demonstrate that this method can be applied to biologically relevant signals including clinical EEG recordings obtained from seizure patients. Finally, we discuss how this method can be applied to study ISA to identify and assess the seizure onset zone.

## INTRODUCTION

Epilepsy is one of the most prevalent neurological conditions affecing an estimated 3.4 million people in the United States [1]. A third of patients with epilepsy continue to experience seizures despite medication treatment [2]. Although many of these patients pursue surgical options, 40-70% of surgical patients continue having seizures after resective surgery [3]. Surgical intervention is thought to fail in these cases due to the incomplete removal of the culprit brain tissue, a putative and theoretical area known as the epileptogenic zone (EZ). Because the EZ is an area that can only be defined post-surgically, the seizure onset zone (SOZ) is used as a proxy for the EZ during surgical planning [4]. The limited efficacy of surgical interventions suggests that our ability to identify the SOZ/EZ is insufficient. Thus, identifying features that are unique to the SOZ/EZ has great potential for improving surgical outcomes.

Low frequency ictal activity is a biomarker that has recently become of interest in the characterization of seizure recordings. Recent data from a number of studies suggest that the EZ may be identified by the presence of very low frequency oscillations [5-7] (for a review, see Lee et al. (2020)). Termed “infraslow activity” (ISA) or direct current (DC) shifts in the literature, this low frequency band is typically defined as activity below 0.1 or 0.5Hz. This frequency band is much slower than the 1-70Hz band at which clinical EEGs are typically interpreted. The study of low frequency activity in seizure recordings, however, has been hampered by technical limitations. EEG systems necessarily include a high-pass filter early in the measurement chain to remove large artifactual drifts that can saturate recording elements such as the amplifier. This filter unfortunately attenuates legitimately seizure-related low frequencies, making ISA difficult to study in clinical EEG recordings. Although a few studies have utilized DC amplifiers to try to bypass this issue [9-12], DC amplifiers are not immune to saturation issues (Supplementary Fig. S1). Furthermore, standard clinical EEG equipment does not utilize DC amplifiers, making observing ISA directly in clinical recordings difficult. Consequently, development of a method to evaluate ISA in recordings obtained with alternating-current (AC) amplifiers is desirable.

In this study, we present a novel approach to digitally reconstruct attenuated low frequency activity in clinical EEG recordings using a deconvolution-based inverse filter. First, we show that a clinical EEG recording system’s unit impulse response (UIR) may be derived by differentiation of the system’s step response. We then use this UIR to deconvolve a variety of synthetic signals to demonstrate successful restoration of attenuated low frequencies. We then show that this method is stable when applied to clinical intracranial recordings obtained from temporal lobe epilepsy patients. Finally, we discuss how this method may be applied to study ISA as it relates to the identification and assessment of the SOZ.

## RESULTS

In linear time invariant (LTI) systems, the effects of recording elements such as filters on signals may be mathematically described as a convolution of the input signal with the system’s characteristic unit impulse response (UIR) (Fig. 1). Inversely, the input of this system can be found by deconvolution of the output signal with the UIR (Fig. 1). If the LTI element in question is a filter, the deconvolution operation may be thought of as an “inverse filter” that allows for recovery of frequencies attenuated by the filter. While this concept is mathematically straightforward, the challenge of applying this method to real-world signals lies in the ability to accurately characterize the UIR of the recording system.

**Figure 1.**
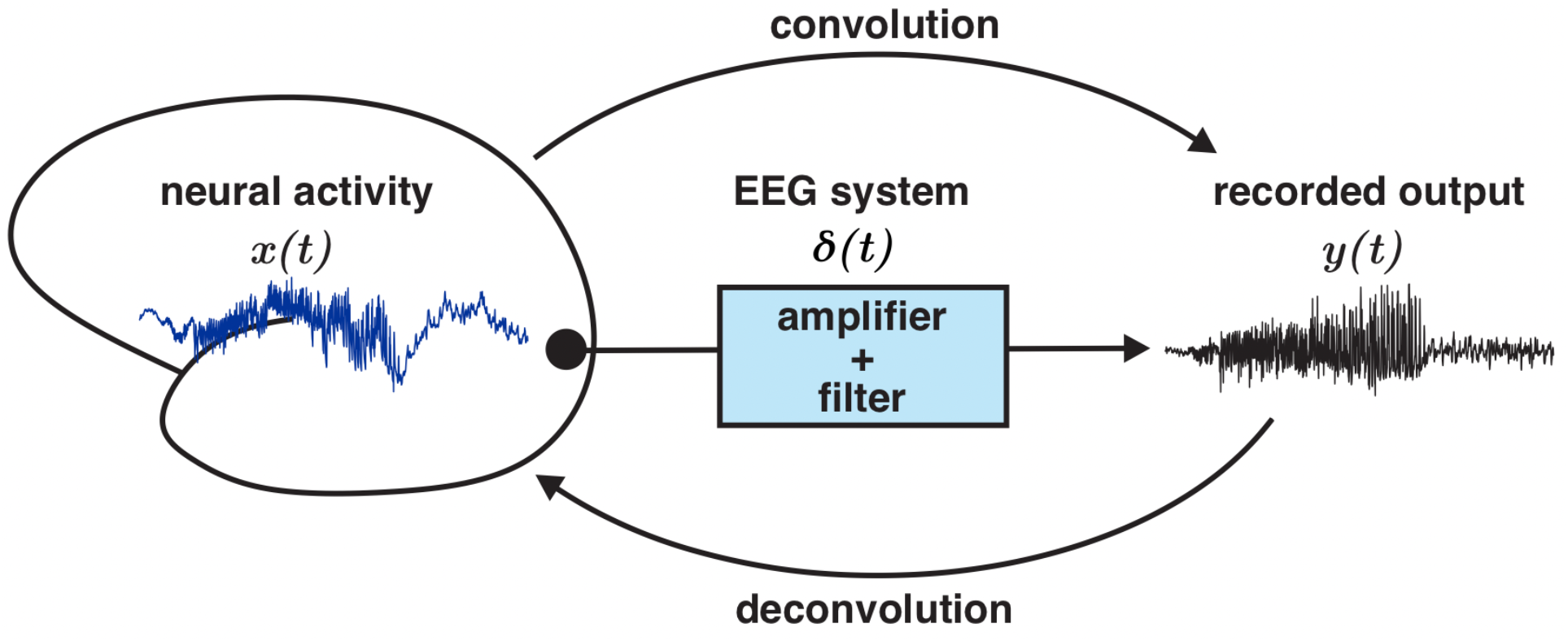
Elements of a linear time invariant (LTI) system may be described by convolution and deconvolution operations. In a LTI system, output function *y(t)* may be described by a convolution of the input signal *x(t)* with the system’s unit impulse response *δ(t)*. Inversely, the input signal may be obtained by deconvolving *y(t)* with *δ(t)*. The unit impulse response fully characterizes the LTI system in the time domain. In this schematic, the LTI system is the EEG recording machinery that includes elements such as amplifiers and high-pass filters.

### Characterization of the unit impulse response of a clinical EEG system

Mathematically, the UIR of a system is the derivative of the system’s step response [13]. To characterize the UIR of a clinical EEG unit (Natus XLTEK Brain monitor with Connex headbox), we used a digital/analog (D/A) converter to input a synthetically generated step function (Fig. 2A) (Methods). The output measured by the EEG system (i.e., the system’s step response) was fitted with a 9^th^ order polynomial (Fig. 2B, C). Taking the derivative of this polynomial approximation resulted in the putative characteristic UIR the recording system (Fig. 2D). The accuracy of this UIR was verified by deconvolving the recorded output of a test function with known low frequency activity. The test function was a series of two step functions (Fig. 3A). Deconvolution of the recorded output of these step functions (Fig. 3B) with the UIR resulted in a reconstruction that resembled the input (Fig. 3C). Notably, the flat feature of the step functions was restored, confirming that this method is appropriate for reconstructing DC shifts. The deconvolution operation was also robust to noise, as the square shape of the reconstruction was preserved even with the addition of high levels of normally distributed random noise (Fig. 3D-F).

**Figure 2.**
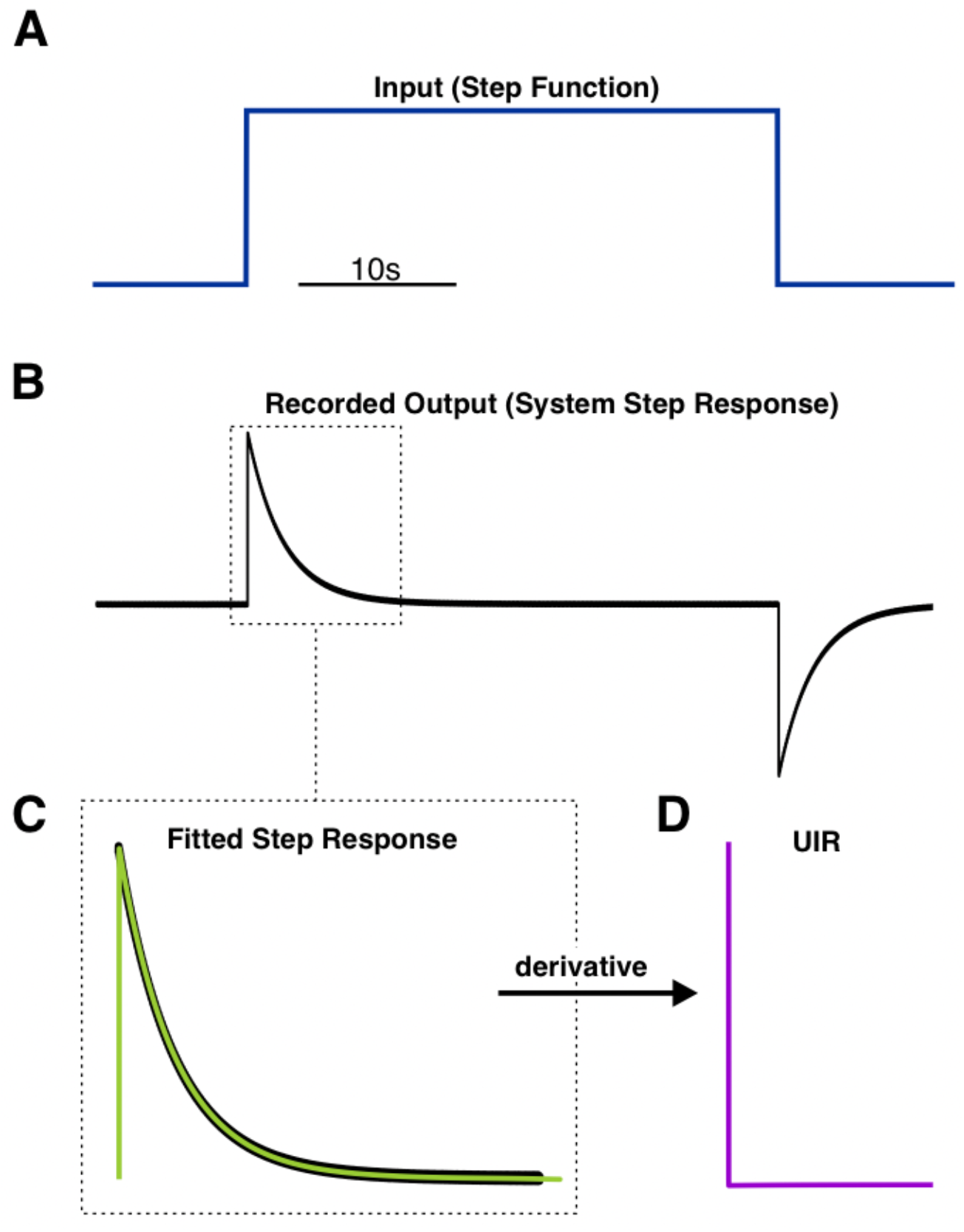
Determination of an EEG system’s unit impulse response (UIR) by measuring the system’s step response. A synthetically generated step function was used as input into the EEG system **(A)**. The measured output was the system’s step response **(B)**. This step response was fitted with a 9^th^ order polynomial (light green trace) **(C)**. Taking the derivative of this polynomial function resulted in the system’s UIR **(D)**.

**Figure 3.**
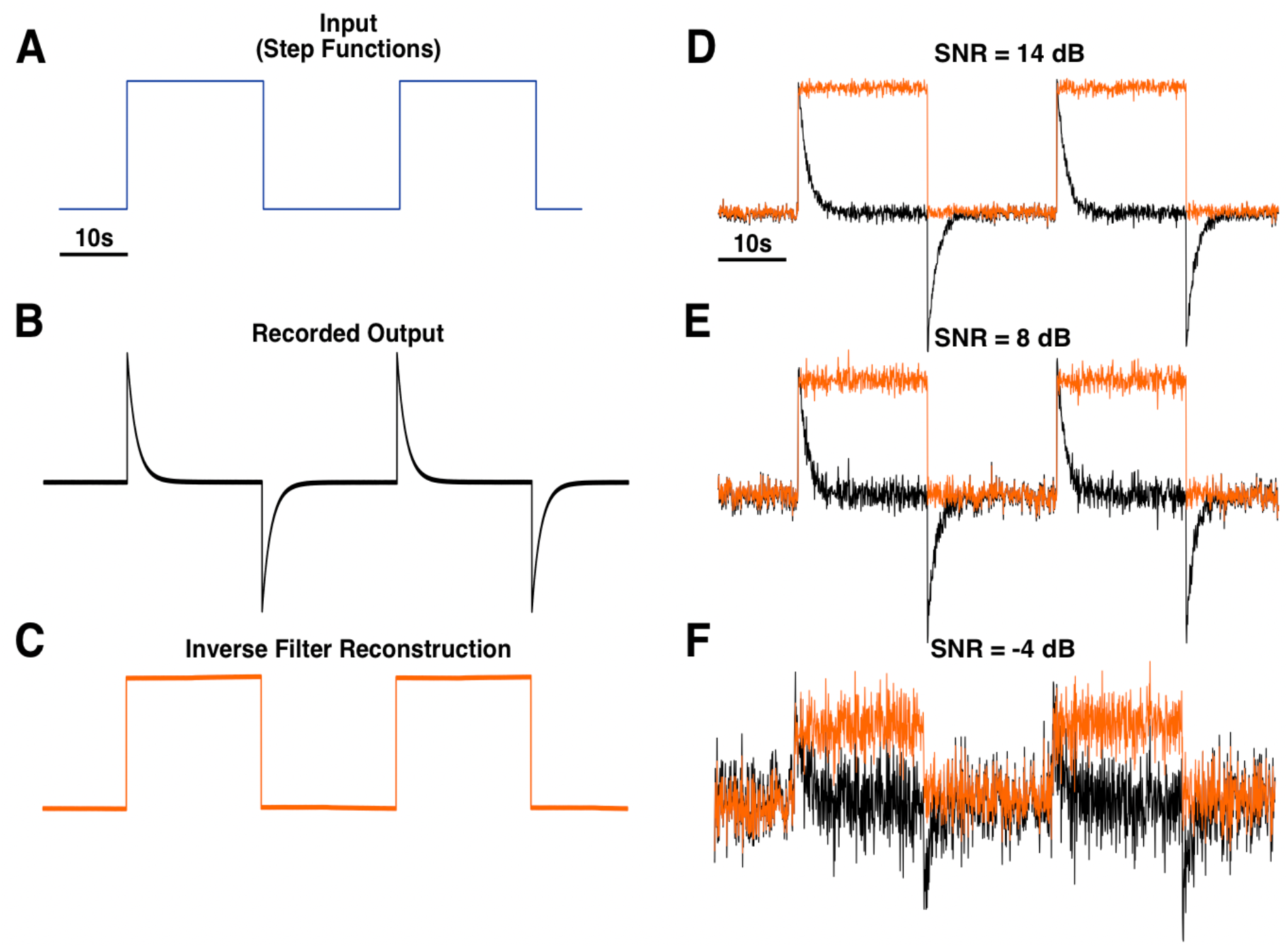
Application of deconvolution-based inverse filter to step functions. A series of two step functions was used as a test input signal **(A)**. The EEG system’s recorded output **(B)** was deconvolved by the unit impulse response (UIR) depicted in Figure 2D, which resulted in the reconstruction of the original step function **(C)**. This method was able to reconstruct the general step function shape with high fidelity even with the addition of varying levels of normally distributed random noise **(D, E, F)**. In panels D, E, and F, the black traces show the recorded output with added noise, and the orange traces show the inverse filtered reconstructions. SNR = signal-to-noise ratio.

### Validation of deconvolution-based inverse filter using synthetic signals with known low frequency components

We then tested our inverse filter procedure on two different types of synthetic signals. All inputs were generated digitally in MATLAB and converted to an analog signal using a digital/analog converter to be used as inputs into the EEG system (Methods, Supplementary Fig. S2). The first test signal was a step function with a 10Hz sine overlay (Fig. 4A). As expected, the DC component of the recorded output was greatly attenuated by the system’s 0.1Hz high-pass filter while the 10Hz sine was unaffected (Fig. 4B). Deconvolution of this output with the UIR reconstructed a function that closely resembled the original synthetic input (Fig. 4C).

**Figure 4.**
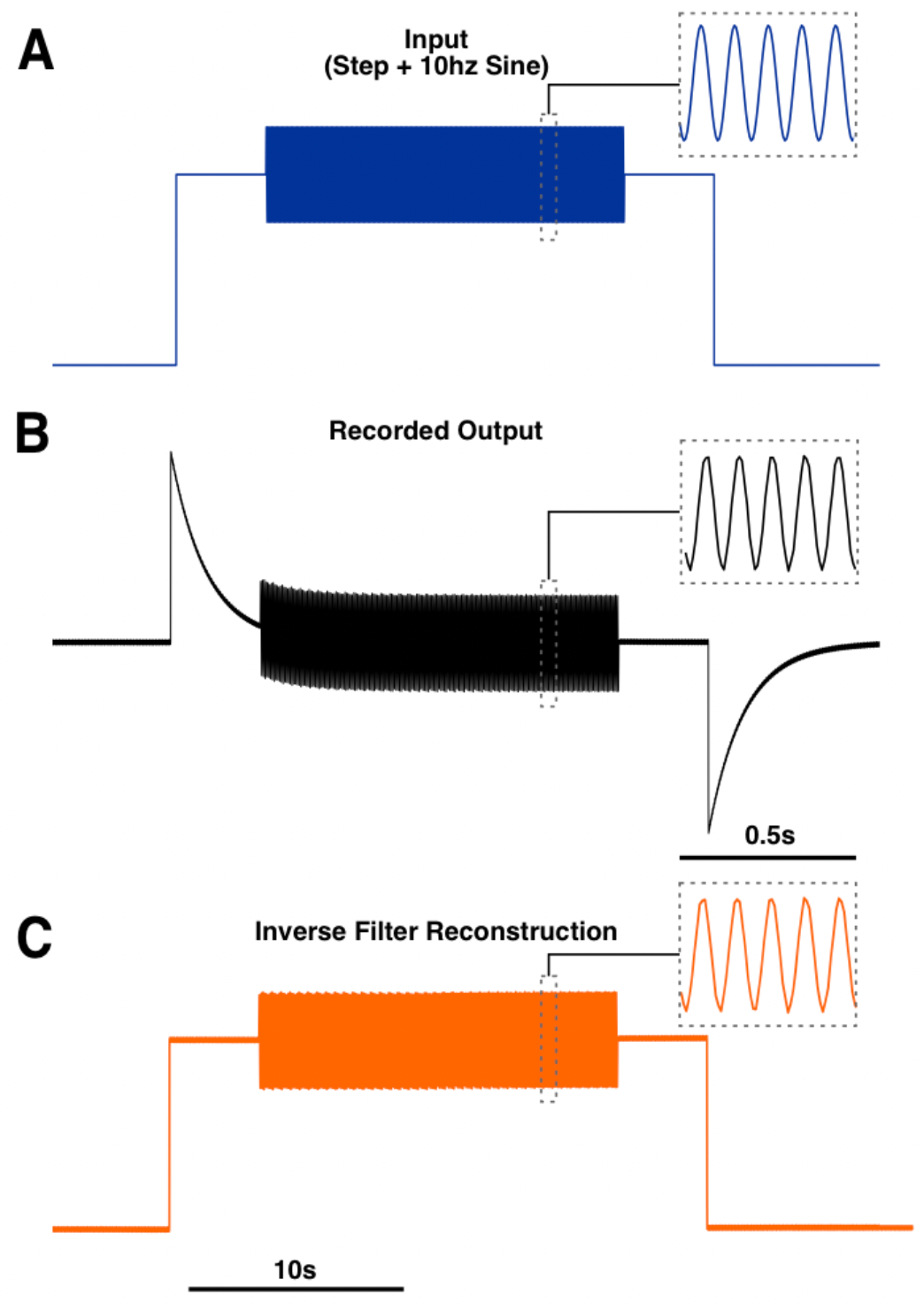
Inverse filter reconstruction of a step function with a 10Hz oscillation. A signal composed of a 10Hz oscillation riding on the top of a step function was used as input into the EEG system **(A)**. The system’s recorded output **(B)** was deconvolved with the system’s UIR, resulting in a reconstruction of the original step function **(C)**. The dotted line insets indicate a 0.5s window showing the 10Hz oscillation.

Next, we tested an input that was a mixed sine function composed of the following frequencies: 0.01, 0.05, 0.07, 0.15, 0.2, 1, 6, and 10Hz (Fig. 5A1). Presence of these frequency components were confirmed with an amplitude spectrum (Fig. 5A2). Low frequency components were visibly attenuated in the EEG system’s recorded output, and an amplitude spectrum confirmed this attenuation (Fig. 5B1, B2). Inverse filtering of the recorded output resulted in a mixed sine wave with the low frequency components restored, which was evident by visual inspection of the time series and confirmed by the amplitude spectra (Fig. 5C1, C2).

**Figure 5.**
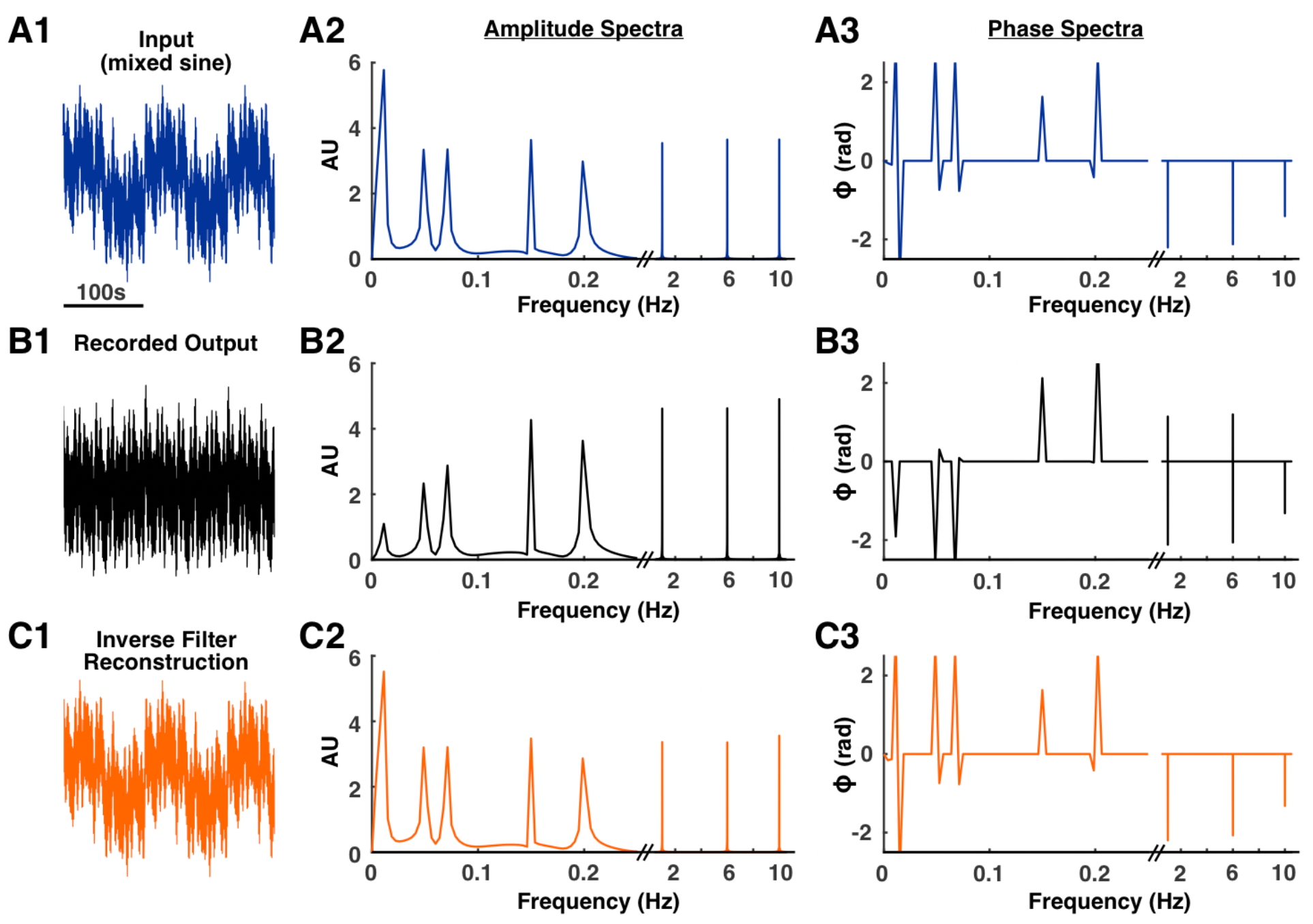
Inverse filter reconstruction of a mixed sine signal. An input signal with known low frequency components was generated by mixing sine waves of varying frequencies **(A1)**. The frequency composition was confirmed with an amplitude spectrum **(A2)**. As expected, activity in frequencies below 0.1Hz was attenuated in the recorded output **(B1, B2)**. The recording process also introduced a phase distortion **(A3, B3)**. Deconvolution of the recorded output with the system’s UIR resulted in a signal with the low frequency components restored **(C1, C2)**. This deconvolution process also corrected the phase distortion **(A3, C3)**.

The phase spectra of the recorded output showed that the recording process distorts the phase of the input signal (Fig. A3, B3). The phase spectra of the original input and reconstruction were similar, demonstrating that the inverse filter was also able to correct the phase distortion caused by the EEG system (Fig. A3, C3). The fidelity of the reconstruction to the original input was quantified by correlation analysis. The time series, amplitude spectra, and phase spectra of the original input were more highly correlated with the reconstruction than the recorded output (Supplementary Table S1).

### Application of deconvolution-based inverse filter to a known biological signal

Next, we wished to test a signal that was of biological relevance. A recording of a hippocampal seizure in a mouse that was available through a public repository was used [14] (Methods). This seizure was originally recorded with a DC amplifier, which allowed preservation and recording of low frequency components. Notably, there is a DC shift prior to the start of the seizure and spreading depolarization activity after the seizure (Fig. 6A1). These features were predictably attenuated in the recorded output (Fig. 6B1). Inverse filtering of this recorded output resulted in a signal that was similar to the original input signal in time, frequency, and phase (Fig. 6C1, C2; Supplementary Fig. S3). This fidelity was confirmed with correlation analysis that showed that for all measures, the input signal was more highly correlated with the reconstruction than the recorded output (Supplementary Table S2).

**Figure 6.**
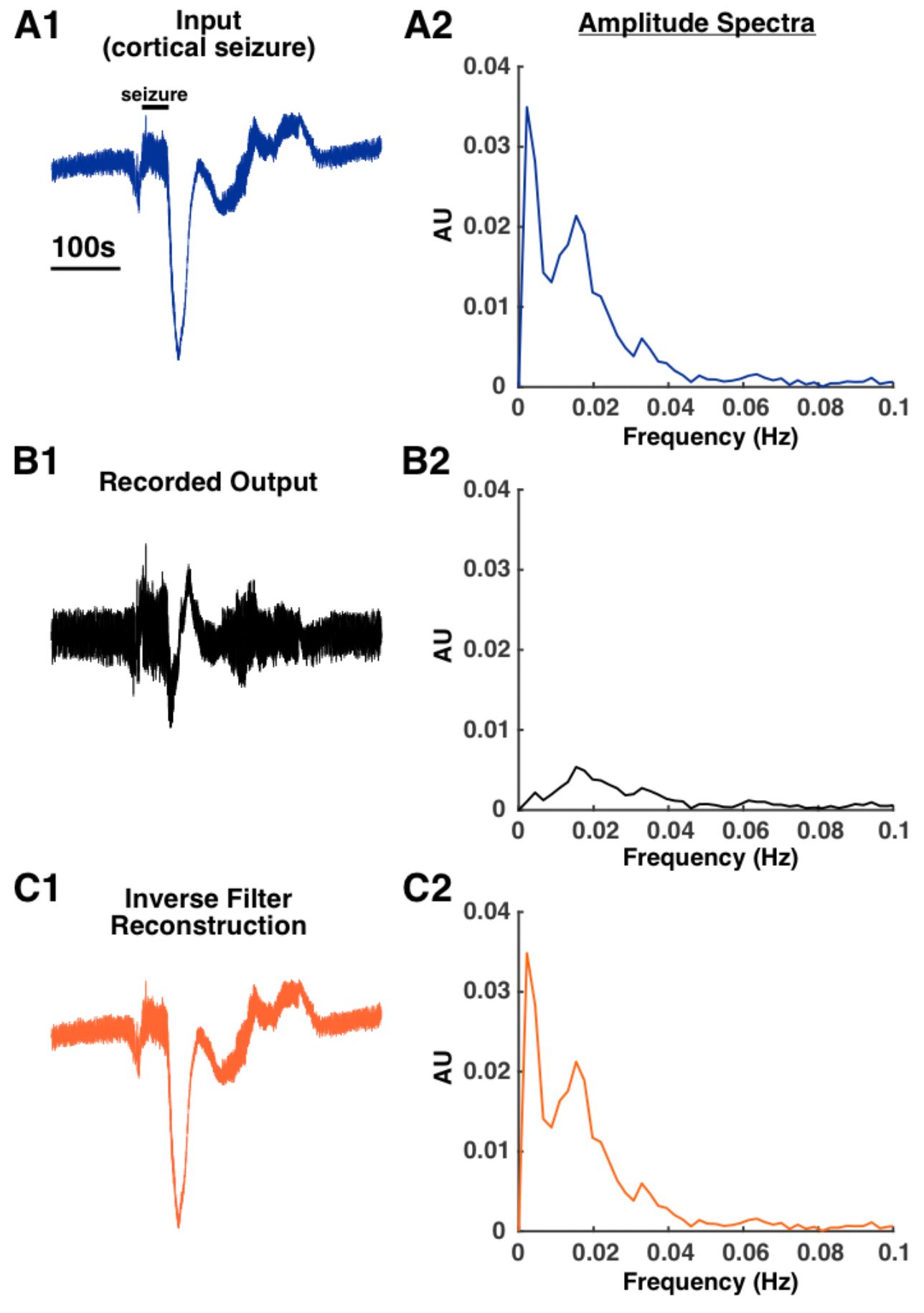
Inverse filter reconstruction of a mouse hippocampal seizure. A recording of mouse hippocampal seizure with prominent low frequency activity shortly after seizure offset was used as the input test signal **(A1, A2)**. This low frequency activity was greatly attenuated in the recorded output **(B1, B2)**. Deconvolution of this recorded output with the system’s UIR resulted in a reconstruction of the original signal with the low frequency components restored **(C1, C2)**.

### Application of deconvolution-based inverse filter to clinical EEG recordings

In the results presented so far, we validated the ability of our inverse filter algorithm to restore low frequencies by using known input signals. Next, we wished to test our inverse filter on a dataset where the ground truth (i.e., the original signal) is unknown. To do this, we used the inverse filter to study low frequency activity in seizure recordings obtained during long-term monitoring of temporal lobe epilepsy patients. Because the EEG set-ups used in the inpatient unit are not portable, characterizing the UIR using the laboratory-based D/A signal generation system is impractical. Therefore, we wished to develop a more convenient method to characterize the UIR. To do this, we took advantage of the fact that the clinical system includes a square wave calibration signal. Because a square wave is simply a series of step functions, we recorded this calibration output, averaged three step responses, and fitted a function to this average to approximate the system’s step response (Supplementary Fig. S4A). To check that the UIR was characterized correctly, this UIR was used to deconvolve the recorded calibration signal, which resulted in a function resembling a square wave (Supplementary Fig. S4B).

The UIR derived from the calibration signal was used to deconvolve intracranial recordings obtained from epilepsy patients undergoing presurgical monitoring for medically intractable temporal lobe epilepsy (Supplementary Table S3). Figure 7 shows example reconstructions of right-sided hippocampal depth (RHD) electrodes for two different patients. The depth probe is inserted along the anterior-poster axis of the hippocampus, with RHD1 being the most anterior electrode (Methods).

**Figure 7.**
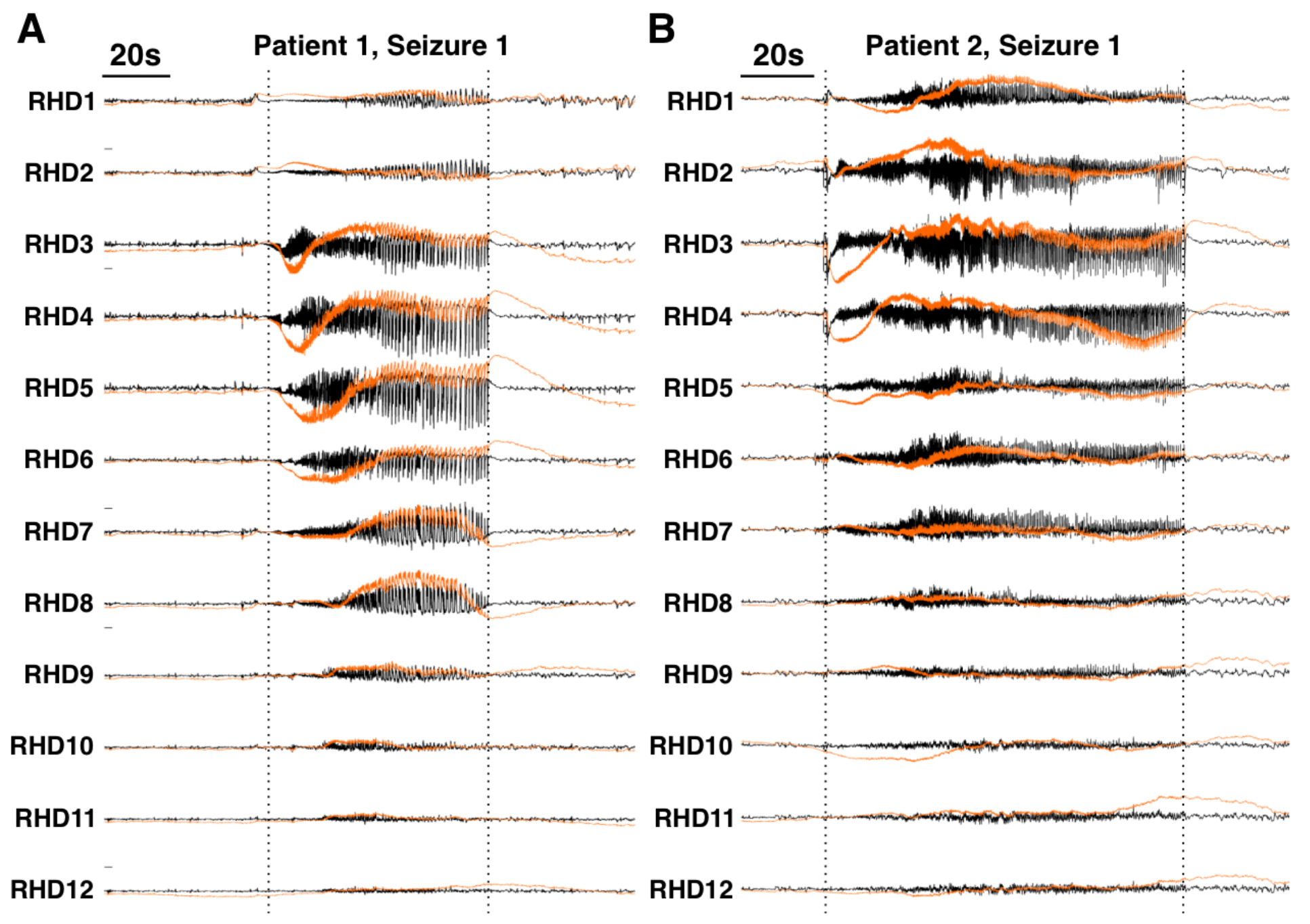
Inverse filter reconstruction of intracranial ictal recordings for two temporal lobe epilepsy patients. Black traces are the raw recordings and orange traces are the inverse filtered signals. In both patients, large amplitude, low frequency shifts are present at the start of the seizure. RHD = right hippocampal depth. Vertical dotted lines indicate seizure onsets and offsets.

To demonstrate that the reconstructions successfully recovered low frequency signals, high resolution time-frequency analysis was performed using a Morlet wavelet. An example of the wavelet analysis for a single channel result for Patient 1, Seizure 1 is shown in Fig. 8. A distinct lack of power in frequencies below 0.1 Hz was observed in the analysis of the raw signal (Fig. 8B), as expected by the presence of the 0.1Hz high-pass filter in the recording system. The inverse filtered reconstruction showed a clear increase in power in the sub-0.1Hz frequency band that is not present in the raw signal (Fig. 8C). The increase in power was also temporally aligned with the seizure onset (vertical dotted lines).

**Figure 8.**
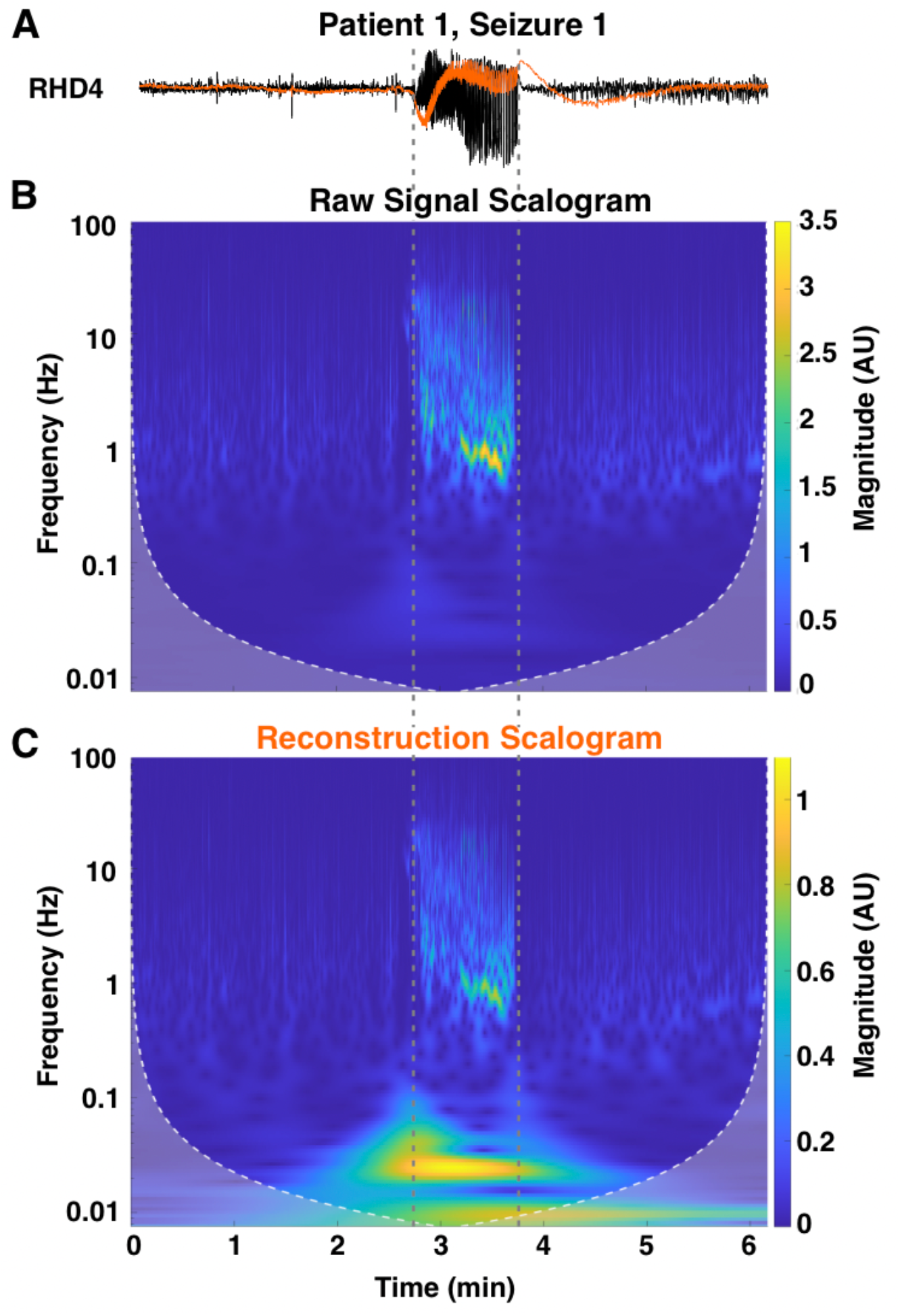
Wavelet analysis of channel RHD4 for the recording for Patient 1, Seizure 1. The inverse filtered reconstruction (orange trace) shows a large low frequency component at seizure onset that is not apparent in the raw recording (black trace) **(A)**. Time-frequency analysis confirmed that frequencies below 0.1Hz are severely attenuated in the raw recording **(B)** but restored in the reconstruction **(C)**. The vertical dotted lines indicate seizure onset and offset.

The restoration of these low frequency components allowed for observations not readily apparent in the raw recordings. In Patient 1, channels RHD3 and RHD4 showed prominent downward shifts at the start of the seizure (Fig. 7A). Channel RHD3 also showed a small upward shift prior to seizure onset. Notably, channels RHD3 and RHD4 were identified as the seizure onset zone (SOZ) during the presurgical assessment of this patient. The SOZ was determined by visually identifying channels that first showed ictal activity in the conventional clinical band of 1-70Hz. Channels RHD5 and RHD6 also showed prominent downward shifts that occurred shortly after the shifts in RHD3 and RHD4. Patient 2 showed similar patterns except the downwards shifts were more concentrated in channels RHD3 and RHD4 (Fig. 7B). For Patient 2, RHD1-8 were identified as the SOZ during conventional clinical assessment. For both patients, these patterns were replicated in a second seizure recording (Supplementary Fig. S5), showing that this inverse filter method performs reliably and consistently when applied to clinical recordings.

## DISCUSSION

### Advantages of the deconvolution-based inverse filter

The deconvolution-based inverse filter presented in this study offers several advantages over current approaches to studying infraslow activity (ISA) in clinical recordings. The use of DC amplifiers has been suggested as a way to preserve low frequency signals in EEG recordings, and some studies have successfully used such set-ups to study ISA [9-12]. This method, however, has not been a practical solution for studying ISA in larger clinical datasets because DC amplifiers are not used in standard clinical equipment. Furthermore, even DC amplifiers have amplifier range limitations that are determined by the power supply potentials. When signals exceed this range, signals become clipped and useful information is lost (Supplementary Fig. S1). One distinct advantage of the method presented in this study is the ability to apply this technique retroactively to clinical recordings obtained with an AC amplifier. The method is entirely computational and does not require any specialty equipment. Consequently, the inverse filter algorithm can be applied to any recordings that are obtained from a system for which a unit impulse response (UIR) can be successfully characterized.

This method is also able to reconstruct the phase information of the original input signal. Hardware filter components, such as those in EEG systems, introduce phase distortions during the filtering process. Our inverse filter method not only reconstructs the attenuated frequency components but also restores the original phase relationships across the spectrum (Fig. 5A3, B3, C3; Supplementary Fig. S3). The fidelity of the phase spectrum is important as it allows for accurate comparisons of the relative timing of signal components. This timing is particularly important when studying the activity of different frequency bands in relation to the seizure onset or when studying relationships such as ISA-HFO coupling.

Some other studies have posed the idea of an inverse filter to reconstruct low frequency activity in EEG recordings [15, 16]. The inverse filter algorithms presented in these studies, however, necessitate characterization of various filter parameters such as resistance and capacitance. This requires knowledge of the precise specifications of the EEG system’s real-time high-pass filter implementation, which may not be readily available, proprietary, or otherwise difficult to obtain. In contrast, the method presented in this study is completely non-parametric, and no specific attributes of the equipment need to be known to characterize the system’s UIR. Furthermore, the UIR may be obtained for any clinical recording equipment that include a native calibration signal, which allows this method to be potentially applied more ubiquitously to clinical recordings (Supplementary Fig. S4).

### Limitations of the deconvolution-based inverse filter

The deconvolution-based inverse filter relies completely on the ability to accurately characterize the UIR of the recording system. Since the UIR is specific to the model of the amplifier, the UIR must be re-characterized if a different model of amplifier is used. Consequently, any updates to hospital recording systems that involves equipment changes will require a recharacterization of the UIR.

This method can be computationally intensive as the computation time scales exponentially with the length of signal being reconstructed. Depending on the computing resources available, inverse filtering longer signals (> 4hrs) may be not practical, although this issue may be bypassed by reconstructing a series of shorter clips and concatenating the results. Significant downsampling of the signals can be also used to reduce computing time since high sampling rates are not necessary to study low frequency activity.

Finally, this method will also reconstruct any low frequency artifacts that can obscure or confound seizure-related ISA. Therefore, any signal being used must be reasonably noise-free, and reconstructions should be screened for results that likely contain artifactual drifts.

### Potential clinical applications

The most distinct advantage of this deconvolution-based inverse filter is that it may be applied retroactively to clinical recordings of seizures. Because ISA has emerged as a topic of interest in identifying epileptogenic tissue (reviewed in Lee et al. (2020)), the ability to observe low frequency activity in larger clinical datasets is critical for further investigation. Although the exact clinical utility of ISA and its potential mechanistic role in seizure generation is beyond the scope of this study, the clinical examples presented here suggest that distinct patterns of ISA can be observed in the SOZ. For Patient 1, the clinically defined SOZ were channels RHD3 and RHD4. Interestingly, these two channels also showed the most prominent ISA at the seizure onset (Fig. 7A). This is consistent with literature that have suggested that ISA is concordant with the SOZ [17, 18]. Large amplitude, downward ISAs were also observed in channels RHD5-6, but they occurred after the ISA in channels RHD3-4 and had a longer time course. The mechanistic implication of these observations is a target for future studies, but our current results suggest that ISA patterns allows for differentiation between electrodes in a way that is not evident by conventional methods.

For Patient 2, the standard clinical assessment identified a much more diffuse SOZ spanning RHD1 to RHD8. In contrast to this assessment, Patient 2 shows a concentration of ISA power in a smaller range of electrodes, namely electrodes RHD3 and RHD4 (Fig. 7B). This observation is consistent with the literature that have suggested that ISA may allow delineation of a smaller epileptogenic area compared to conventional methods [19-22].

## METHODS

### Recording of synthetic and biological test signals

The three synthetic signals (step, step with 10Hz sine overlay, mixed sine) were generated in MATLAB (MATLAB, Natick, MA, USA) (Fig. 3A, 4A, 5A1). Signals were generated with a sampling rate of 1000 samples/s. The mouse hippocampal seizure recording was downloaded from a publicly available repository (https://doi.org/10.5281/zenodo.5655535) [14] in HDF5 format, downsampled to 1000 samples/s, and converted to a *.mat file (Fig. 6A1). These signals were saved as text files, and a plain-text word processor (WordPad, Microsoft, Windows 10) was used to add a header and convert them into *.atf files that could be read by Clampex (Molecular Devices LLC, San Jose,CA, USA, v.10.4.1.10).

Next, a Clampex episodic stimulation protocol was used with a digital/analog (D/A) converter (Digidata 144A, Molecular Devices LLC, San Jose, CA, USA) to generate an analog output corresponding to each test signal. The output from the D/A converter was attenuated by a factor of 1/1000 using a 10 kOhm/10 Ohm resistor pair in series. This attenuated signal was recorded with a clinical EEG machine (Natus XLTEK Brain monitor with Connex headbox). The recorded signals were exported in the *.edf format for subsequent analysis in MATLAB.

### Unit impulse response curve fitting and deconvolution

Curve fitting to approximate the step response was performed using the *polyfit* interface in MATLAB. A 9^th^ order polynomial was used. Deconvolution was performed using the *deconv()* function. Signals were zeroed by subtracting the mean prior to deconvolution. MATLAB scripts that demonstrate this process step-by-step are available in a Github repository (https://github.com/sominlee14/deconvolution_based_inverse_filter).

### Signal processing and statistical analysis

All signal processing and statistical analyses were performed in MATLAB. All reconstructed signals were filtered with a 2^nd^ order high-pass Butterworth filter with a cutoff frequency of 0.005Hz prior to visualization. This filter step necessary because the deconvolution process in some cases introduces a triangular drift due to the accumulation of rounding errors. The frequency of this drift is entirely dependent on the epoch length of the signal being deconvolved and is equal to 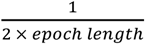. To avoid confounding this triangular drift with ictal-associated slow activity, we only used signals with lengths at least three times the period of the lowest frequency of interest. For example, to analyze frequencies around 0.005Hz, the signal being inverse filtered was at least 600 seconds long.

### Patients and clinical data acquisition

Clinical EEG recordings were collected from two patients undergoing phase II monitoring for medically intractable right temporal lobe epilepsy at the University of Chicago Adult Epilepsy Center (Supplementary Table S3). Written informed consent was obtained through a process approved by The University of Chicago Institutional Review Board. For both patients, intracranial recordings were collected using depth electrodes placed into the right hippocampus, 12 channel depth electrodes placed along the length of the right hippocampus with contact #1 (RHD1) being most anterior and contact #12 (RHD12) being most posterior. Other intracranial electrodes were placed in other locations, but recordings from these channels were not used for this study.

All patient recordings used were collected using an XLTEK EEG recording system (Natus Neurolink IP EEG Amplifier, INBOX-1166A/B, Natus, Pleasanton, CA, USA). Signals were digitized at 1024 samples/s and referenced to the FCz electrode. The raw broadband (0.1-344Hz) was converted into *.mat files using a custom C++ routine. Signals were downsampled to 256 samples/s for all subsequent analyses.

## Data Availability

All scripts and data except human EEG recordings are available at https://github.com/sominlee14/deconvolution_based_inverse_filter

## ACKNOWLEDGEMENTS

We thank Dr. Naoum Issa, and Graham Smith for valuable discussions and feedback. S.L. and W.v.D were supported by NIH Grant R01 NS-084142. S.L. was supported by The University of Chicago MSTP Training Grant T32GM007281. A.K.T. was supported by the Comer Children’s Development Board (Race for Kids at Comer).

## AUTHOR CONTRIBUTIONS

Research Design: S.L., S.W., W.v.D.

Data collection: S.L., J.H., A.K.T., Y.E., P.W., S.W., W.v.D.

Data analysis: S.L., W.v.D.

Manuscript writing & editing: S.L., A.K.T., S.W., W.v.D.

## DATA AVAILABILITY

With the exception of patient EEG recordings, all input signals, recordings, and custom scripts used in this study can be found at https://github.com/sominlee14/deconvolution_based_inverse_filter. Patient recordings are not publicly available due to HIPAA protections, but they can be made available upon reasonable request to the corresponding author (W.v.D), provided that all data sharing follow protocols compliant with HIPAA policies.

