## Supplemental Materials for "Digital reconstruction of infraslow activity in human intracranial ictal recordings using a deconvolution-based inverse filter"

### Supplementary Figures

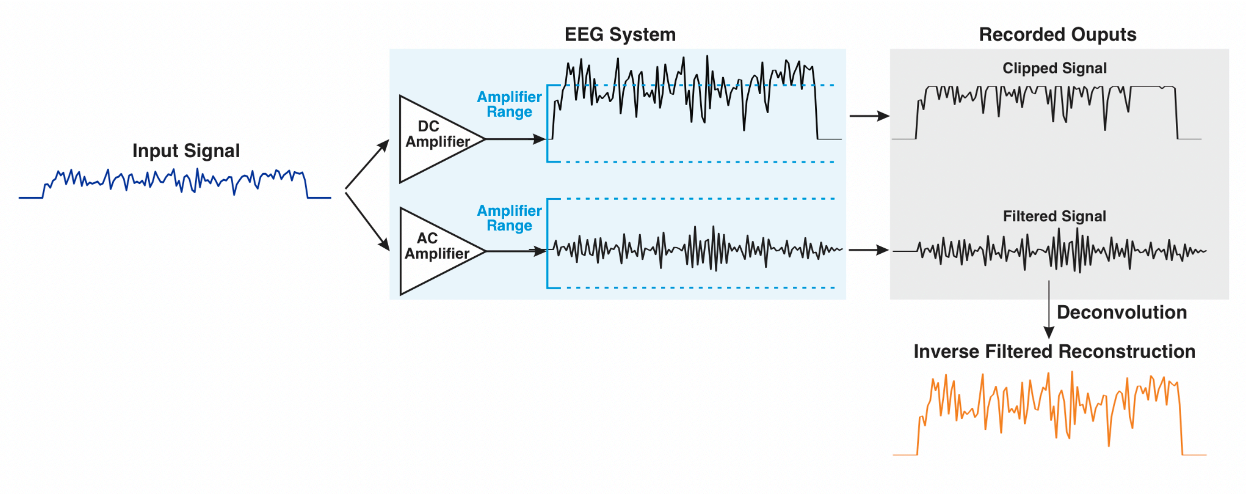


**Supplementary Figure S1.** Limitations in using direct-current (DC) and alternating-current (AC) amplifiers in recording low frequency activity. Although the use of a DC amplifier allows preservation of low frequency components, signals can become clipped if the large amplitude slow activity exceeds the range of the amplifier (top chain). AC amplifiers include a high-pass filter that allows signals to comfortably stay within the amplifier range, but low frequency activity is greatly attenuated (lower chain). Using a deconvolution-based inverse filter algorithm to digitally restore low frequency activity in signals measured with an AC amplifier is one solution to this issue (bottom, orange trace).

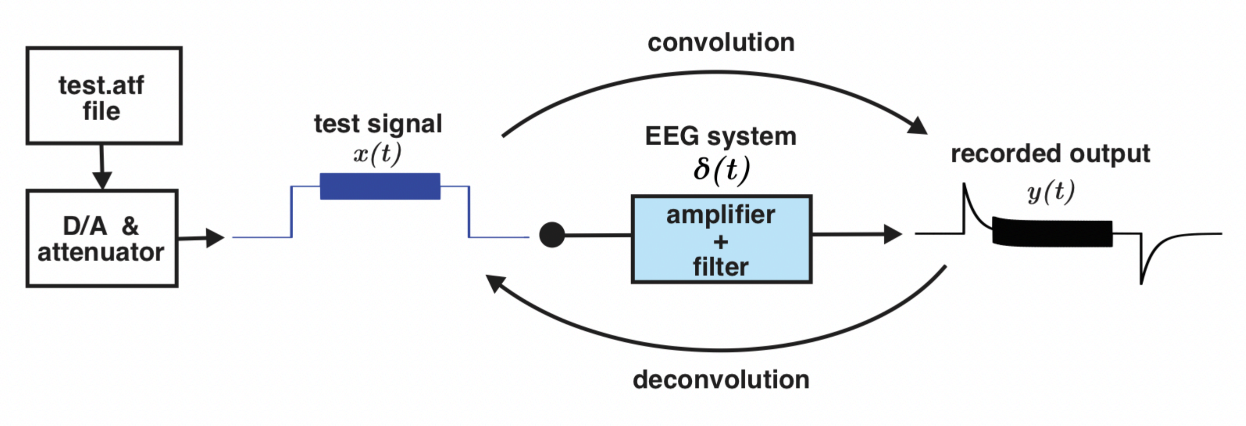


**Supplementary Figure S2.** Schematic of digital-to-analog (D/A) conversion of digitally-generated synthetic signals to use as inputs into an EEG system. An *.atf file containing the digital version of the test signal was read by Clampex software and converted into an analog signal using a D/A converter. The output from the D/A converter was attenuated before being recorded by a clinical EEG machine. Due to the high-pass filter that is part of the EEG system, low frequencies are attenuated in the recorded output. This recorded output then can be deconvolved to return the original test signal.

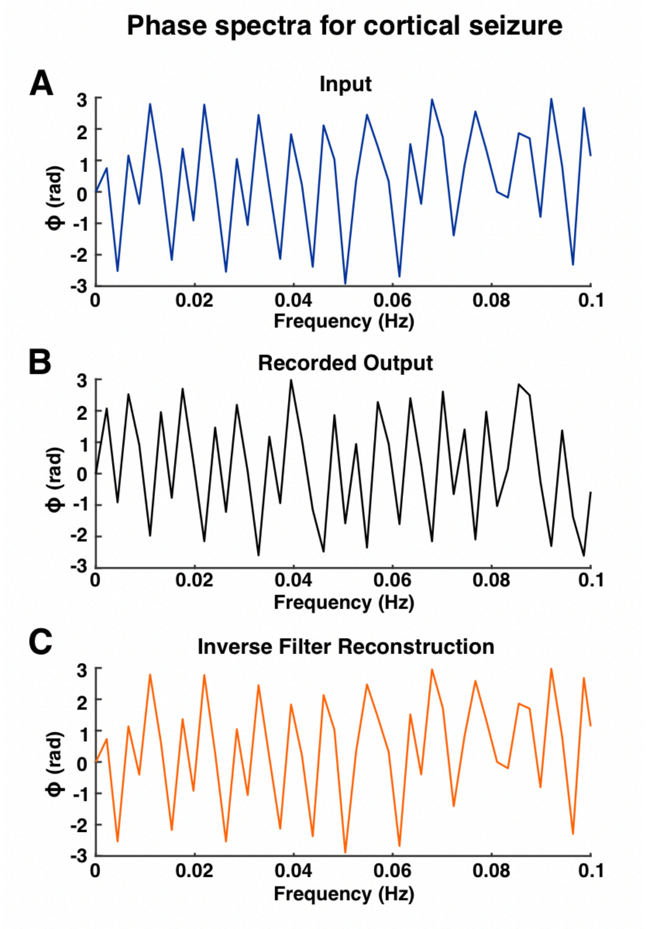


**Supplementary Figure S3.** Phase spectra for the mouse hippocampal seizure recording and inverse filter reconstruction shown in Figure 7. The phase spectrum for the input signal **(A)** is different from the phase spectrum for the recorded output **(B)** because the recording machinery and associated filter introduces phase shifts. The deconvolution-based inverse filter reverts these phase shifts **(C)**.

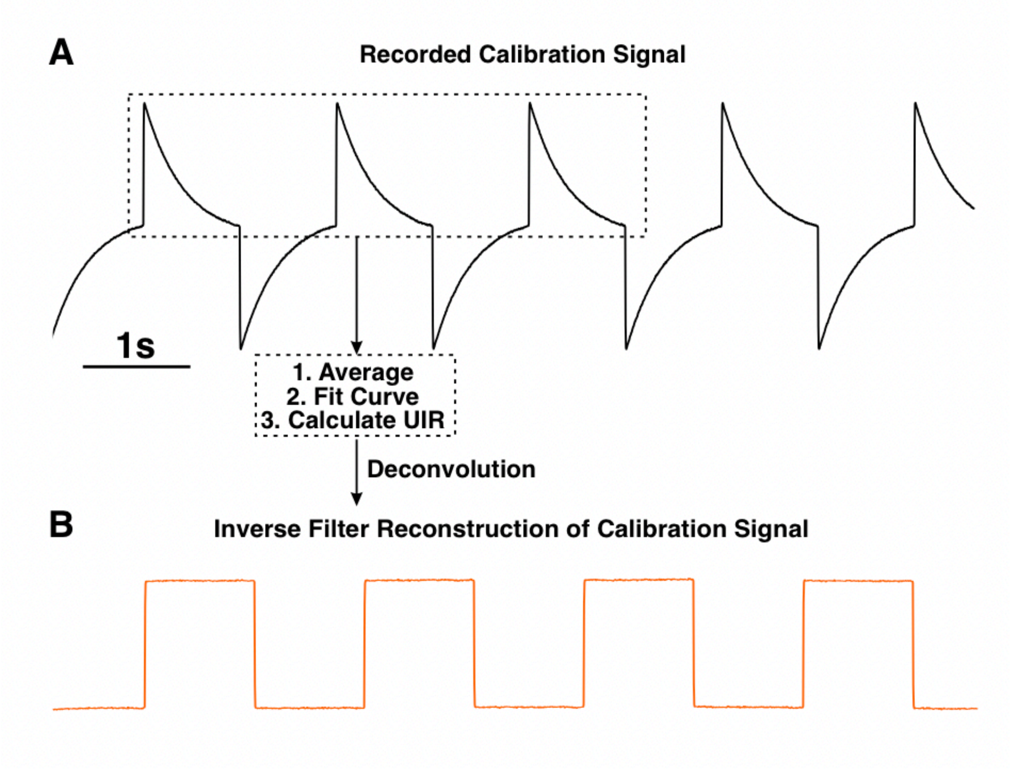


**Supplementary Figure S4.** Characterizing the unit impulse response (UIR) for an inpatient XLTEK EEG system using the native calibration signal. The native calibration signal is known to be a square wave, which may be thought of as a series of step functions. The average of three peaks in the calibration signal was used to approximate the system's step response and calculate the UIR **(A)**. Deconvolution of the calibration signal with the calculated UIR resulted in the expected square wave **(B)**.

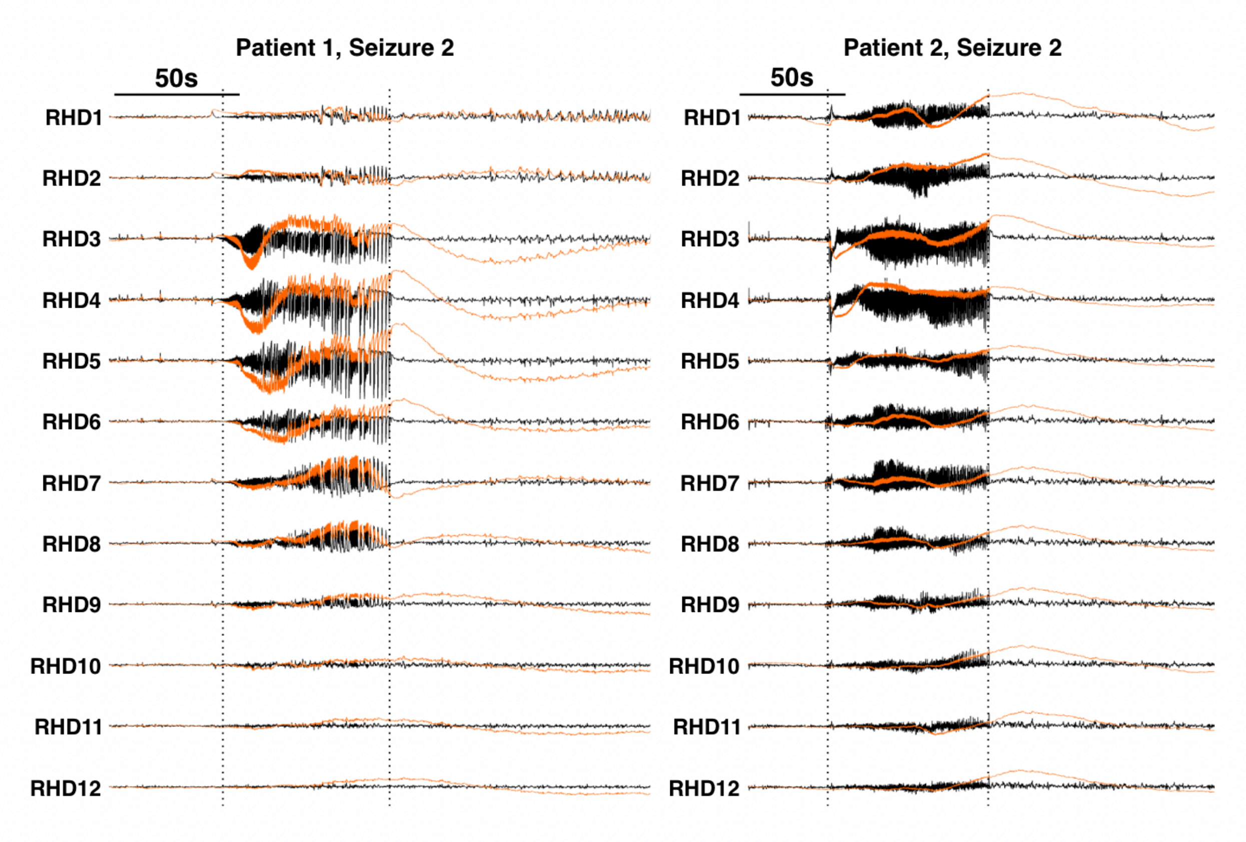


**Supplementary Figure S5.** Additional examples of inverse filter reconstructions of ictal recordings from Patient 1 and Patient 2. Black traces are the raw recordings, and the orange traces are the inverse filter reconstructions. The low frequency activity patterns observed in Seizure 1 for both patients were replicated in a second seizure recording. RHD = right hippocampal depth electrode. Vertical dotted lines indicate seizure onsets and offsets.

### Supplementary Tables

**Supplementary Table S1.** Correlation results for comparing the time series, amplitude spectra, and phase spectra for the mixed sine signal. In all comparisons, the correlation between the input and reconstruction was higher than the correlation between the input and recorded output.

Correlation results for mixed sine signal

*Time series correlations*

| | | Recorded output | Inverse filtered reconstruction |
|---|---|---|---|
| Input | Pearson correlation | r = 0.70 | r > 0.99 |
| | Sig. two-tailed | p < 0.001 | p < 0.001 |
| | N | n = 68304 | n = 68304 |

*Amplitude spectra correlations*

| | | Recorded output | Inverse filtered reconstruction |
|---|---|---|---|
| Input | Pearson correlation | r = 0.84 | r > 0.99 |
| | Sig. two-tailed | p < 0.001 | p < 0.001 |
| | N | n = 34153 | n = 34153 |

*Phase spectra correlations*

| | | Recorded output | Inverse filtered reconstruction |
|---|---|---|---|
| Input | Pearson correlation | r = 0.036 | r > 0.99 |
| | Sig. two-tailed | p < 0.001 | p < 0.001 |
| | N | n = 34153 | n = 34153 |

**Supplementary Table S2.** Correlation results for comparing the time series, amplitude spectra, and phase spectra for the mouse cortical seizure. In all comparisons, the correlation between the input and reconstruction was higher than the correlation between the input and recorded output.

Correlation results for mouse cortical seizure

*Time series correlations*

| | | Recorded output | Inverse filtered reconstruction |
|---|---|---|---|
| Input | Pearson correlation | $r = 0.22$ | $r > 0.99$ |
| | Sig. two-tailed | $p < 0.001$ | $p < 0.001$ |
| | N | $n = 116797$ | $n = 116797$ |

*Amplitude spectra correlations*

| | | Recorded output | Inverse filtered reconstruction |
|---|---|---|---|
| Input | Pearson correlation | $r = 0.66$ | $r > 0.99$ |
| | Sig. two-tailed | $p < 0.001$ | $p < 0.001$ |
| | N | $n = 58399$ | $n = 58399$ |

*Phase spectra correlations*

| | | Recorded output | Inverse filtered reconstruction |
|---|---|---|---|
| Input | Pearson correlation | $r = 0.72$ | $r = 0.90$ |
| | Sig. two-tailed | $p < 0.001$ | $p < 0.001$ |
| | N | $n = 58399$ | $n = 58399$ |

**Supplementary Table S3.** Patient demographics and diagnoses. Age refers to age at which the recording was obtained. TLE = temporal lobe epilepsy

| Patient # | Age (years) | Diagnosis | Engel score |
|---|---|---|---|
| 1 | 20's | Drug-resistant TLE | II |
| 2 | 50's | Drug-resistant TLE | I |
